# Differences in sepsis from skin and soft tissue infections among patients with vs. without mental health disorders

**DOI:** 10.64898/2026.10.31.26366979

**Authors:** Prajna Sahadeo, Sandhya LoGalbo, Evan Wu, Alessandro Drudi, Illan Saji, Hannah Oliva, Aliyah Traore, Daimia Foster, Jeffrey Wang, Vinayak Sahadeo, Mark Richman

**Author notes:** The HLPRS (Hofstra Longitudinal Pre-medical Introduction to Research Society) Cohorts.

## Abstract

**Introduction:** Sepsis is a severe health condition that requires rapid and attentive treatment. The rise in sepsis cases reveals a lack of immediate supportive care in the healthcare system. This lapse may be attributed to a delay in sepsis recognition, patient incompatibility with care plan, and conditions that make treatment more difficult. Building on previous research, this paper seeks to explore the correlation between patients with mental health conditions and sepsis development. State how we define quality of sepsis care.

**Method:** Data from 2018-2019 from our ED sepsis quality database was analyzed using T-tests, comparing the characteristics, conditions, and quality of sepsis care of dermatologically-septic patients with and without mental health conditions.

**Results:** Between January 1, 2018 and December 31, 2019, 584 LIJ ED patients had sepsis that met inclusion criteria for quality review. Of these, 57 (9.8%) had sepsis of dermatologic origin, of which 30 (52.6%) were female (p = 0.4318). The source of sepsis was the lower extremity in 36.8% of mental health patients vs. 68.4% of non-mental health patients (p = 0.045) and sacral decubitus ulcer in 36.8% of mental health patients vs. 5.3% of non-mental health patients (p = 0.004). In neither univariate nor multivariate analysis were there statistically-significant differences in outcomes in patients with vs. without a mental health issue. However, the need for mechanical ventilation trended lower among patients with mental health disorder (p = 0.141) while the outcome of 3-hour bundle met trended higher among that same group (p = 0.11).

**Discussion:** Compared to patients with mental health issues, those without mental health issues had higher rates of sepsis due to lower extremity infections and sacral decubitus ulcer infections. None of the outcome variables met statistical significance. However, dermatologic sepsis patients without mental health issues tended to be more sick than those with mental health issues, as gauged by rates of septic shock and ICU admission (both trending toward statistical significance). They also trended toward receiving lower quality care, as measured by adherence with the 3-hour sepsis bundle. Failure to achieve statistical significance might be due to the small sample size. This is likely the case, as, for example, there was a substantial difference in the percent of multi-racial patients between those with (5.3%) vs. without (28.9%) mental health illness, yet even this large difference did not meet statistical significance (p = 0.104).

## Introduction

Sepsis is a life-threatening ailment caused by an exaggerated immune response to infection that results from the body’s response to infection, which causes injury to its own tissues and organs. Sepsis is a major health problem with increasing prevalence, high costs, and poor outcomes. In the United States, sepsis accounts for about 5.2% of hospital expenditures, more than $20 billion per year, and hospitalizations for more than 1 million people. ^1^ Severe sepsis and septic shock are a subset of sepsis conditions. This subset of conditions accounts for 25–50% of in-hospital mortality, ranging from 45% to 65% after six months of discharge.

^2^ Mortality rates rise with sepsis severity: 10-20% for sepsis, 20-40% severe sepsis, and 40-80% for septic shock.^3^ If patients with these conditions survive, they have increasing chronic complications, morbidity, high costs of care, and decreasing quality of life.^4,5^Many of the over 1. million annual cases of sepsis in the United States present to the Emergency Department (ED).^6,7^Improved outcomes, including decreased mortality, are achieved with prompt identification of sepsis and treatment with broad-spectrum antibiotic agents and intravenous fluids,^8^ as recommended in the Surviving Sepsis Campaign and endorsed by clinical practice guidelines and the Centers for Medicare and Medicaid Services (CMS).^9,10^

Per the Surviving Sepsis Campaign, sepsis is defined as having ≥2 SIRS criteria (temperature >38.0°C or hypothermia <36.0°C, heart rate >90 beats/minute, respiratory rate >20 breaths/minute, leukocytosis >12*10^9^/L or leukopenia <4*10^9^/L) and a likely bacterial source of their SIRS. Patients meeting these criteria should, within 3 hours of sepsis being suspected, have: 1) Serum lactate measured, 2) For patients with elevated lactate or hypotension (mean arterial pressure (MAP <65), intravenous fluid (IVF) bolus of at least 30 mL/kg administered, unless contraindicated by evidence or risk of volume overload, 3) In fluid-non-responsive patients with elevated lactate or hypotension, giving vasopressors to raise blood pressure, 4) Blood cultures drawn before antibiotics given, 5) Broad-spectrum antibiotics administered after blood cultures drawn, 6) For patients with elevated serum lactate, a repeat serum lactate and re-assessment of the need for additional IVF within 6 hours of sepsis being suspected, to determine whether tissue perfusion improved with initial IVF and vasopressors.^10^

Patients may fail to receive the complete bundle for a variety of reasons. There may be a delay in recognizing sepsis. Patients may have a contraindication to the full 30 mK/kg IVF bolus, or an advanced directive limiting use of bundle interventions. They might: 1) be currently psychotic or manic and unable to explain their symptoms appropriately, 2) not understand their condition or the care they’re offered, 2) refuse care, 3) be agitated or aggressive, or 4) take medications whose side effects could be misinterpreted as sepsis (eg, neuroleptic malignant syndrome (NMS)).

Previous research has demonstrated patients with severe mental illness and cardiovascular disease or diabetes are 30% less likely to receive recommended cardiovascular or diabetes care, including hospitalization, diagnostic tests, appropriate medications, and invasive procedures.^11^ It is possible such disparities apply to severe sepsis or septic shock, as well. However, very few such studies have been performed. A national cross-sectional study in Sweden found patients with severe mental health disorders had increased risk of death associated with influenza/pneumonia (OR = 2.06, 95% CI [1.87–2.27]) and sepsis (OR = 1.61, 95% CI [1.38–1.89]). They also had an increased risk of hospitalization associated with influenza/pneumonia (OR = 2.12, 95% CI [2.03–2.20]) and sepsis (OR = 1.89, 95% CI [1.75– 2.03]).^12^

Our previous research^13^ found that, compared with national rates of mental illness, a higher percent of septic patients (26.9% vs. 21.0%) had mental illness (mostly neurocognitive). Septic patients with mental illnesses tended to be older, from a psychiatric facility/group home, have Medicare, have at least one comorbidity, and have a respiratory source of sepsis (while patients without mental illness more-often had a urinary tract source of sepsis). Patients without mental illnesses more-often had immune-modifying medications and malignancy. There was no difference in rates of diabetes. The higher rate of respiratory source of sepsis among those with mental illnesses may be due to a higher risk for aspiration or contagion of respiratory pathogens in group homes or psychiatric facilities.^14^

The primary aim of this current study is to characterize patients whose cause of sepsis is from a skin and soft tissue (SSTI) dermatologic source (eg, cellulitis, abscess, wound infection), comparing patient and visit attributes for those with a mental health condition vs. those without. We postulate there may be significant differences, as those with mental health conditions might be prone to certain forms of SSTI dermatologic-related sepsis (eg, wound infections for patients with dementia vs. abscesses among those with substance abuse disorders). The secondary aim is to explore the quality and timeliness of sepsis bundle care provided to patients, as indicated by the percent of patients in each group who received 3- and 6-hour bundle-compliant care.

## Methods

Northwell Health is a 28-hospital health system largely operating in Long Island and New York City. Long Island Jewish (LIJ) Medical Center is a 583-bed tertiary care teaching hospital serving an ethnically and socio-economically diverse population. The adult ED sees approximately 100,000 patients per year. Members of LIJ’s Department of Quality Services collect and submit sepsis data to state and federal agencies. Patients with a contraindication to bundle components or an advanced directive are removed from the sepsis database. We extracted data from LIJ from 2018 and 2019.

### Inclusion criteria

1. Location: LIJ ED
2. Dates of patient ED presentation: 1/1/18-12/31/19
3. Sepsis (per LIJ sepsis data extraction and reporting management system)

### Exclusion criteria

1. Pregnant
2. Patients with advanced directives for non-aggressive medical care (these will already have been excluded through the sepsis data extraction and reporting management system)

Patients who met inclusion criteria were stratified into either having, or not having, one of the mental health conditions in Appendix A.

Specifically, we will compare differences among patients with vs. without a mental health condition in:

1. Primary aim

a. Age
b. Gender
c. Patient source of arrival (eg, home vs. nursing home)
d. Comorbidities
e. Type of infection (cellulitis vs. abscess vs. wound infection)
2. Secondary aim

a. 3-hour quality of care (bundle adherence)
b. 6-hour quality of care (bundle adherence)

### Data analysis

Student’s T-test will be used to compare characteristics of dermatologically-septic patients with vs. without mental health conditions, and the proportions who received vs. didn’t receive full sepsis bundle care comparing patients with vs. without acute mental health crises.

This project was deemed to fall under the realm of performance improvement by the Institutional Review Board (IRB #: 24-0076).

## Results

Between January 1, 2018 and December 31, 2019, 584 LIJ ED patients had sepsis that met inclusion criteria for quality review. Of these, 57 (9.8%) had sepsis of dermatologic origin, of which 30 (52.6%) were female (p = 0.4318). There were trends toward differences in the presence of at least one comorbidity, and in racial composition, though none of these reached statistical significance. (**Table 1**),

**table 1.**

| **Variable** | **Mental Health Diagnosis** | **No Mental Health Diagnosis** | **Fisher P value** | **Significance** |
|---|---|---|---|---|
| **Any Comorbidity** | 52.6% | 71.1% | 0.240 | Non-significant |
| **Multi-racial** | 5.3% | 28.9% | 0.104 | Non-significant |
| **Other Asian** | 52.6% | 34.2% | 0.104 | Non-significant |
| **Other Race** | 42.1% | 36.8% | 0.104 | Non-significant |
| **Female Gender (30 female, 27 male)** | 52.6% | 50.3% | 0.4318 | Non-significant |

There were statistically-significant differences in some clinical characteristics of patients with sepsis of a dermatologic origin with a mental health issue vs. those without a mental health issue. The source of sepsis was the lower extremity in 36.8% of mental health patients vs. 68.4% of non-mental health patients (p = 0.045) and sacral decubitus ulcer in 36.8% of mental health patients vs. 5.3% of non-mental health patients (p = 0.004). Wound infections were found in 70.6% of mental health vs. 50.0% of non-mental health patients (trending toward statistical significance with p = 0.240). Culture positive for any organism (staphylococcus, streptococcus, or other organism) was found in 26.3% of MH patients vs. 42.1% of non-MH patients (trending toward statistical significance with p = 0.383). A positive culture source (being either blood, wound, or other) was found in 26.3% of mental health vs. 42.1% of non-mental health patients (trending toward statistical significance with p = 0.382). (**Table 2**)

**table 2.**

| **Variable** | **Mental Health Diagnosis** | **No Mental Health Diagnosis** | **Odds Ratio** | **P-value** |
|---|---|---|---|---|
| **Lower Extremity** | 36.8% | 27.0% | 0.27 | 0.045 |
| **Sacral Decubitus Ulcer** | 36.8% | 5.3% | 10.5 | 0.004 |
| **Cellulitis** | 52.6% | 55.3% | 1.05 | 1 |
| **Abscess** | 15.8% | 15.8% | 1 | 1 |
| **Wound Infection** | 70.6% | 50.0% | 2.4 | 0.24 |
| **Culture Positive (Any Organism)** | 26.3% | 42.1% | 0.49 | 0.383 |
| **Any Positive Source** | 26.3% | 42.1% | 0.53 | 0.382 |

In neither univariate (**Table 3, Figure 1)** nor multivariate analysis (**Table 4**) were there statistically-significant differences in outcomes in patients with vs. without a mental health issue. However, the need for mechanical ventilation trended lower among patients with mental health disorder (p = 0.141) while the outcome of 3-hour bundle met trended higher among that same group (p = 0.11).

**table 3.**

| **Outcome** | **Mental Health Diagnosis (%)** | **No Mental Health Diagnosis (%)** | **Absolute Difference (%)** | **P value** | **Significance** |
|---|---|---|---|---|---|
| **ICU Admission** | 31.60% | 36.80% | -5.3 | 0.775 | Non-significant |
| **In-Hospital Mortality** | 5.30% | 10.50% | -5.3 | 0.655 | Non-significant |
| **Septic Shock** | 36.80% | 47.40% | -10.5 | 0.574 | Non-significant |
| **Mechanical Ventilation** | 10.50% | 26.30% | -15.8 | 0.301 | Non-significant |
| **3-Hour Bundle Met** | 88.90% | 67.60% | 21.3 | 0.11 | Non-significant |
| **6-Hour Bundle Met** | 35.30% | 27.00% | 8.3 | 0.54 | Non-significant |

**table 4.**

| Outcome / Variable | Adjusted OR | Adjusted P | Adjusted Significance | Interpretation |
|---|---|---|---|---|
| **ICU Admission** | 0.75 (0.22–2.58) | 0.646 | Non-significant | No meaningful difference |
| **In-Hospital Mortality** | 0.46 (0.04–4.81) | 0.513 | Non-significant | No meaningful difference |
| **Septic Shock** | 0.81 (0.25–2.66) | 0.727 | Non-significant | Trend: lower in Mental Health |
| **Mechanical Ventilation** | 0.28 (0.05–1.53) | 0.141 | Non-significant | Trend: lower in Mental Health |
| **3-Hour Bundle Met** | 3.99 (0.73–21.78) | 0.11 | Non-significant | Trend: higher in Mental Health |
| **6-Hour Bundle Met** | 1.35 (0.36–5.08) | 0.654 | Non-significant | No meaningful difference |

**fig 1.**
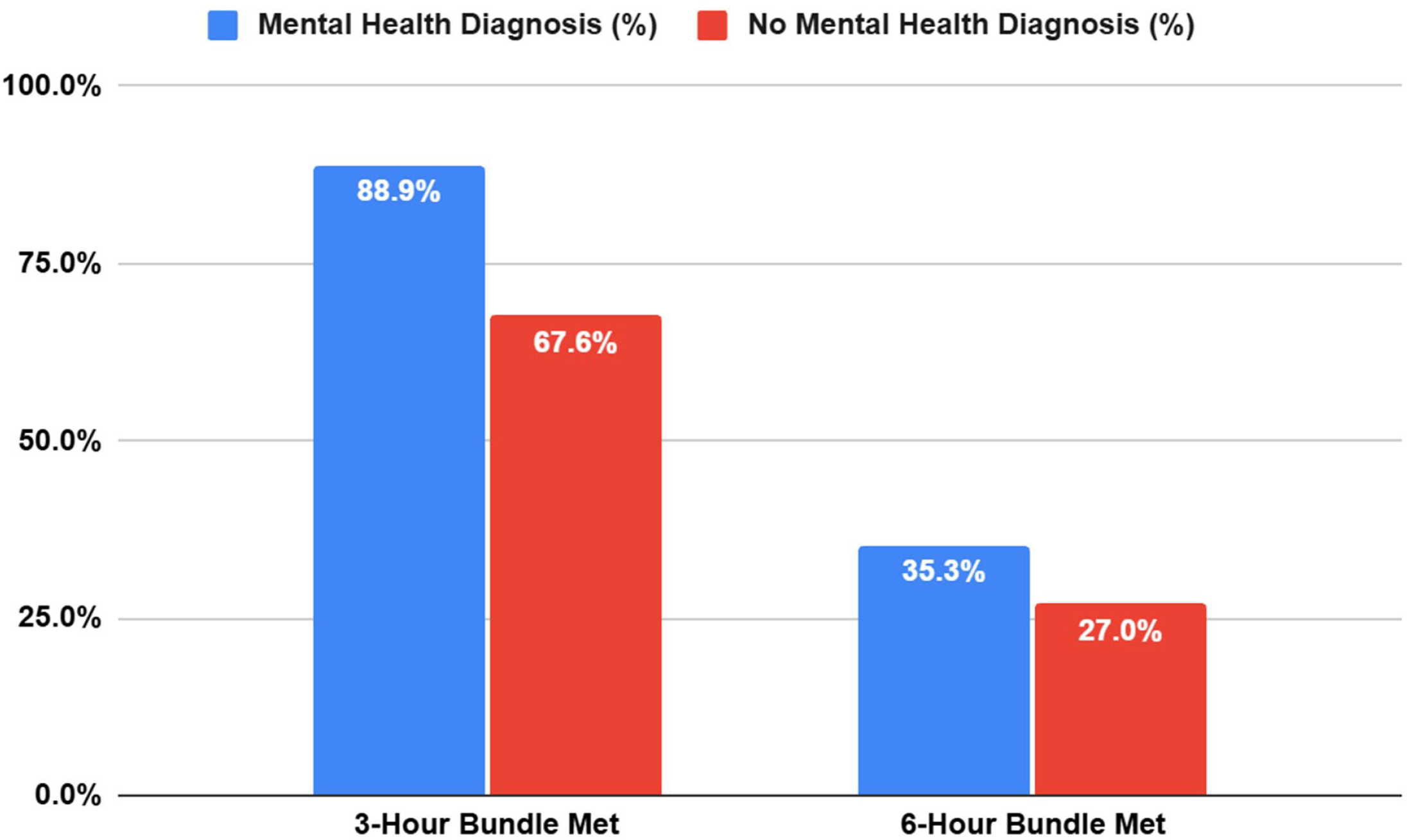

## Discussion

Compared to patients with mental health issues, those without mental health issues had higher rates of sepsis due to lower extremity infections and sacral decubitus ulcer infections. (**Table 2**) None of the outcome variables met statistical significance. (**Table 3**) However, dermatologic sepsis patients without mental health issues tended to be more sick than those with mental health issues, as gauged by rates of septic shock and ICU admission (both trending toward statistical significance). They also trended toward receiving lower quality care, as measured by adherence with the 3-hour sepsis bundle. Failure to achieve statistical significance might be due to the small sample size. This is likely the case, as, for example, there was a substantial difference in the percent of multi-racial patients between those with (5.3%) vs. without (28.9%) mental health illness, yet even this large difference did not meet statistical significance (p = 0.104).

### Limitations

This was a single-site study. Although data was collected from 2018 and 2019, there were only 57 patients with sepsis of a dermatologic origin. Hence, some of the comparisons may not have achieved statistical significance because of a low number of patients.

## Conclusion

Despite previous studies suggesting that patients with mental health conditions receive worse care and result in worse health outcomes,^11,12^ our data shows no significant difference between dermatologic sepsis patient care and the patient’s mental health status. This discrepancy may be due to a low number of patients. Further research with larger patient populations are required to better understand patient care for dermatologic sepsis patients and the effects of patient mental health issues.

## Data Availability

All data produced in the present study are available upon reasonable request to the authors

## Appendix A: ICD-10 codes for severe mental illness

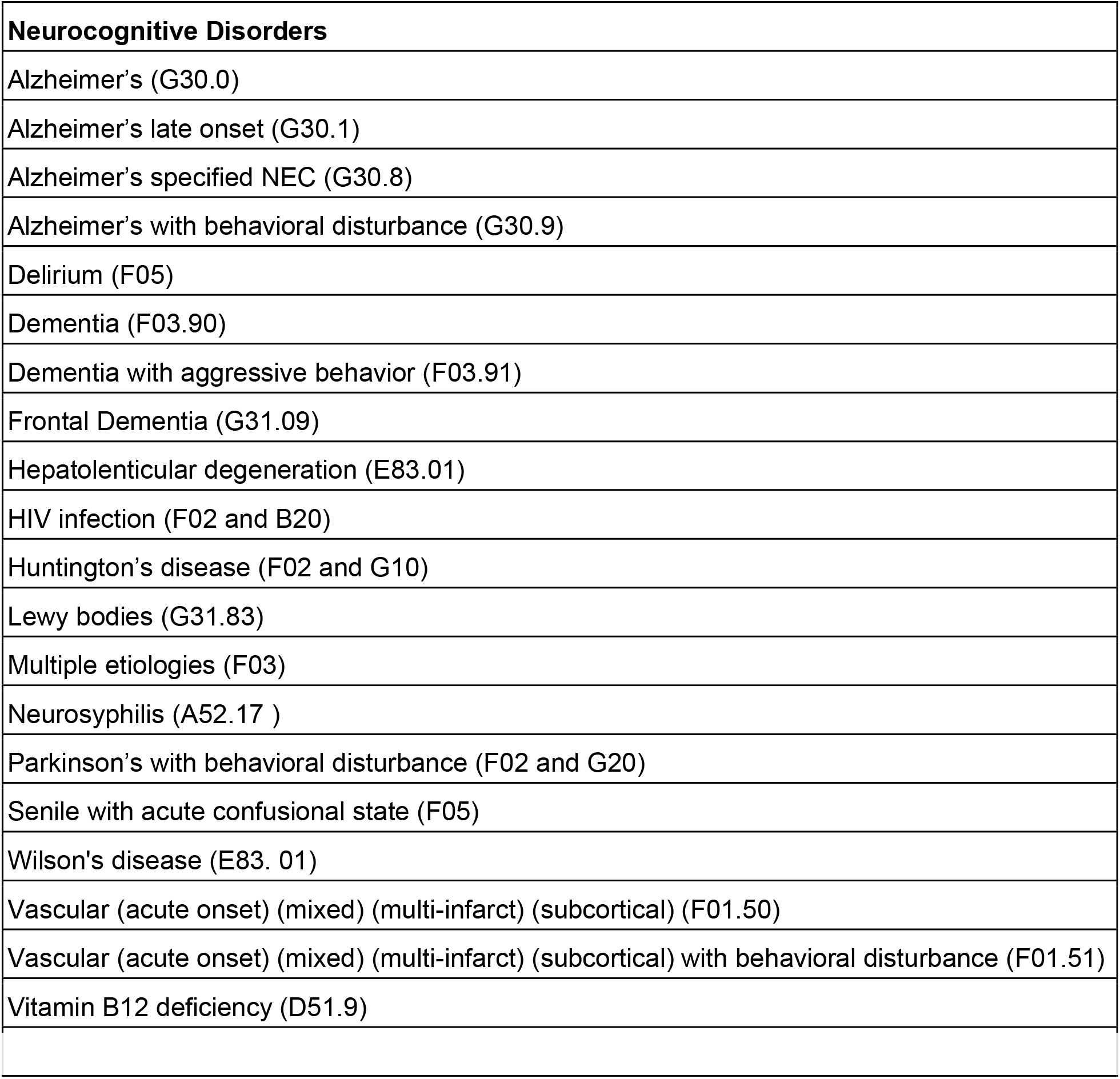
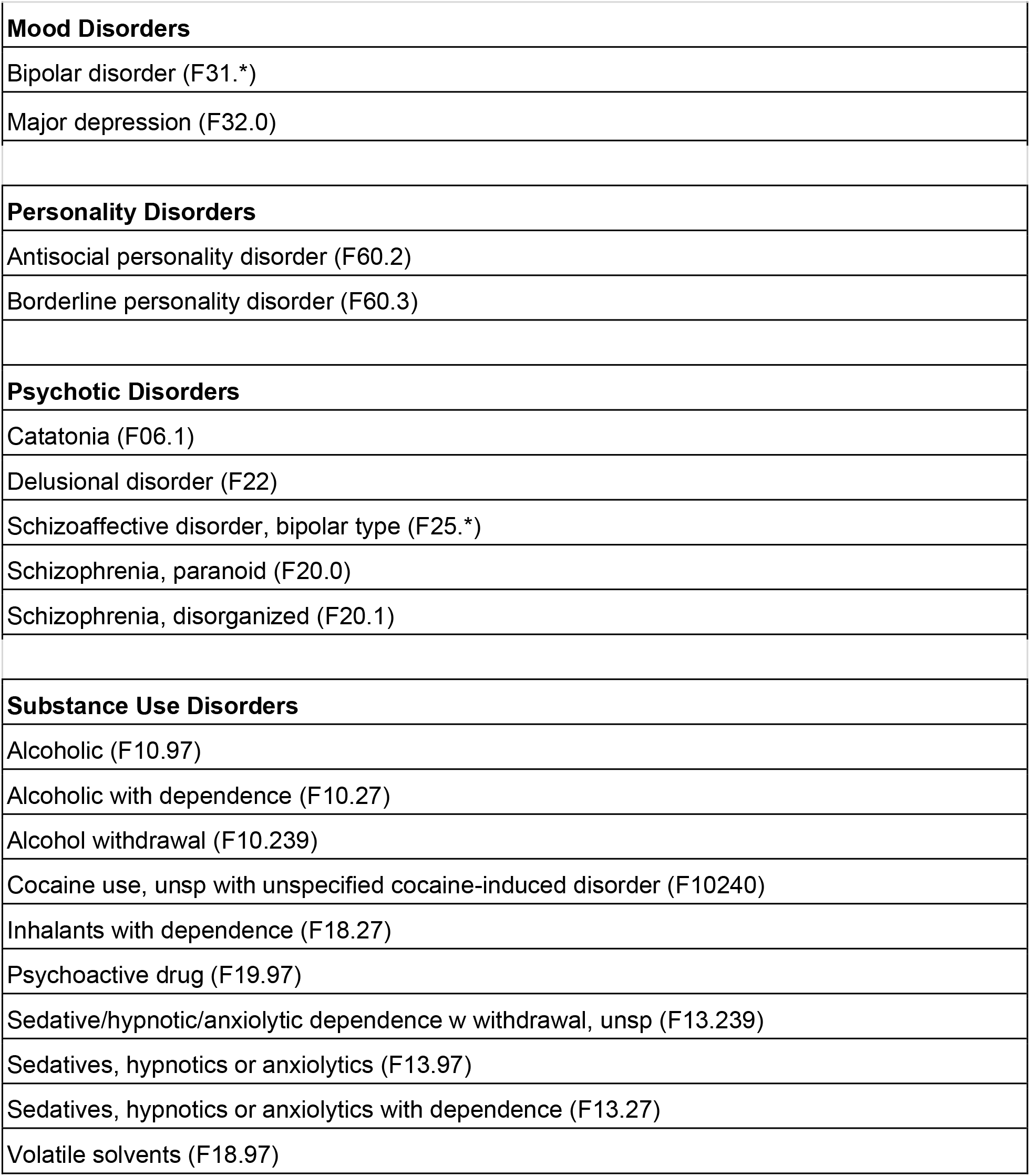

| **Neurocognitive Disorders** |
|---|
| Alzheimer’s (G30.0) |
| Alzheimer’s late onset (G30.1) |
| Alzheimer’s specified NEC (G30.8) |
| Alzheimer’s with behavioral disturbance (G30.9) |
| Delirium (F05) |
| Dementia (F03.90) |
| Dementia with aggressive behavior (F03.91) |
| Frontal Dementia (G31.09) |
| Hepatolenticular degeneration (E83.01) |
| HIV infection (F02 and B20) |
| Huntington’s disease (F02 and G10) |
| Lewy bodies (G31.83) |
| Multiple etiologies (F03) |
| Neurosyphilis (A52.17 ) |
| Parkinson’s with behavioral disturbance (F02 and G20) |
| Senile with acute confusional state (F05) |
| Wilson's disease (E83. 01) |
| Vascular (acute onset) (mixed) (multi-infarct) (subcortical) (F01.50) |
| Vascular (acute onset) (mixed) (multi-infarct) (subcortical) with behavioral disturbance (F01.51) |
| Vitamin B12 deficiency (D51.9) |
| **Mood Disorders** |
| Bipolar disorder (F31.*) |
| Major depression (F32.0) |
| **Personality Disorders** |
| Antisocial personality disorder (F60.2) |
| Borderline personality disorder (F60.3) |
| **Psychotic Disorders** |
| Catatonia (F06.1) |
| Delusional disorder (F22) |
| Schizoaffective disorder, bipolar type (F25.*) |
| Schizophrenia, paranoid (F20.0) |
| Schizophrenia, disorganized (F20.1) |
| **Substance Use Disorders** |
| Alcoholic (F10.97) |
| Alcoholic with dependence (F10.27) |
| Alcohol withdrawal (F10.239) |
| Cocaine use, unsp with unspecified cocaine-induced disorder (F10240) |
| Inhalants with dependence (F18.27) |
| Psychoactive drug (F19.97) |
| Sedative/hypnotic/anxiolytic dependence w withdrawal, unsp (F13.239) |
| Sedatives, hypnotics or anxiolytics (F13.97) |
| Sedatives, hypnotics or anxiolytics with dependence (F13.27) |
| Volatile solvents (F18.97) |

